# Climate Change is expected to expand malaria transmission range and population at risk in Papua New Guinea

**DOI:** 10.1101/2025.06.01.25328753

**Authors:** Stephan Karl, Eloise Skinner, Samuel McEwen, John Keven, Jacob Kisomb, Leanne Robinson, Moses Laman

## Abstract

Rising temperatures can expand malaria transmission into higher altitudes. This is relevant for the projection of future resource needs to sustain malaria control. In Papua New Guinea (PNG), malaria transmission is intense at low altitudes, but new infections are rarely acquired above 1600 m.

The present study applied the basic reproduction number (R_0_) over temperature to assess changes in malaria transmission ranges in PNG over past decades (1960 to 2019) and in the near future by using satellite-based temperature data and climate projections for 2021 to 2040. The analysis considered geographic range shifts for malaria temperature suitability, the resulting changes in the population at risk and effectiveness of interventions.

Malaria temperature suitability ranges have subtly changed between 1960 and 2019, with the proportion of people living in suitable areas increasing from 58% to 61% (equivalent to 249,125 people). Under a conservative climate change model, this proportion is expected increase to 74% by 2040 (equivalent to 2,802,709 people). Interventions had a larger impact on malaria incidence in areas with R_0_<0.3, mitigating the current and future impact of climate change.

Nevertheless, the number of people requiring access to malaria control is expected to double by 2040, to 13.4 million with 2.8 million attributed to climate change alone. The impacted areas are densely populated highlands regions with a more susceptible population and an increased potential for epidemics and clinical disease.

These findings underscore the challenges of climate change for malaria elimination in PNG and highlight the need to accurately guide preparedness and forecast the additional resource requirements.

**Key Findings:**

1. Climate change and population growth are predicted to double the number of people at risk of malaria by 2040
2. The altitude limit for malaria transmission will shift by about 300 m by 2040 as compared to 1960.
3. Current control strategies had a much higher impact to sustainably reduce malaria incidence at higher altitudes

## Introduction

Papua New Guinea (PNG) is the largest Pacific Island Country, and has a fast growing population that is expected to reach 18 million by 2040 [1, 2]. PNG is geographically and ecologically diverse, with thousands of kilometers of coast, expansive low-lying inland areas covered by tropical rainforest, and densely populated highlands regions over 1600 m of altitude. Among other factors such as copious rainfall and mild climate suitable for agriculture, it is likely that the highlands regions of PNG were attractive to early settlers, also for the absence of malaria [3].

PNG reported 1.2 million malaria cases in 2024, but the World Health Organisation (WHO) models suggest over 1.6 million cases, more than 85% of estimated cases in the entire Western Pacific Region [4]. While malaria is a leading cause of illness in the PNG [5], transmission is geographically restricted to areas below approximately 1600 m. Thus, while malaria incidence in lower lying areas can be very high, the PNG Highlands above 1600 m are predominantly malaria-free [6]. As a consequence, populations inhabiting the PNG Highlands (approximately 40% of PNGs total population) do not present extensive acquired immunity, making them potentially more vulnerable to severe manifestations of the disease [7]. Areas between 1200 and 1600 m can also experience large epidemics [8]

Global heating is shifting the natural altitude barrier limiting malaria transmission towards higher altitudes in many geographies across the world [9-12]. In the context of PNG, this may mean a longer transmission season, more frequent epidemics during warmer months, and many more people living in stable transmission environments soon. This implies increased resource requirements for the prevention and control of malaria in and already stretched health systems environment [13]. It is thus important to develop a better understanding of what extent the malaria transmission range will expand or contract in PNG in the next few decades, as this will determine malaria control efforts in the country, and elimination prospects in the for the entire Western Pacific Region, where countries have committed to a malaria elimination agenda until 2030 [14].

The specific aim of the present study was to apply a previously developed temperature R_0_ model [15] to the PNG context to examine the malaria transmission range shifts between 1960 and 2019, and to explore what can be expected in the next two decades until 2040. In addition the present study sought to analyse malaria incidence reduction in relation to R_0_ in the context of currently implemented control interventions for the 2010-2019 decade.

## Methods

Monthly minimum (T_min_) and maximum (T_max_) temperature data was acquired from WorldClim at 2.5 arcmin resolution from Jan 1960 to Dec 2019 (https://www.worldclim.org/)[16]. A future climate prediction for 2021 to 2040 based on the Shared Socioeconomic Pathways 245 (SSP245) model and the ACCESS-CM2 General Circulation Model (GCM) was also acquired from WorldClim (https://www.worldclim.org/) [16]. PNG administrative boundaries level 0, digital elevation (1 arcsec resolution) and population density distribution (30 arcsec resolution) maps were acquired from multiple sources [17-19]. The PNG administrative boundary level 3 (local-level government area) was shared by the PNG National Malaria Control Program. The PNG administrative boundaries layers were overlaid with a 2.5 by 2.5 km grid layer and the grid layer was clipped to the PNG geography using the QGis software (version 3.36.2). Average temperature (monthly T_min_ and T_max_ for 1960 to 2019 and the 2021-2040 prediction), digital elevation and population density zonal statistics were calculated for each grid cell using the QGis software with Orfeo ToolBox [20], and merged into a comma delimited (csv) file. Average temperature was calculated as T_avg_=(T_min_+T_max_)/2.

The relative, temperature-dependent basic reproduction number, R_0_(T), indicating temperature suitability was calculated based on the average temperature for each grid cell and available time point (monthly for 1960-2019 and average monthly for the 2021-2040 period) using the model previously published by Mordecai et al. [15]. The model integrates all components of vector and parasite biology (including biting rate, survival rates, etc.) to estimate R_0_ across temperatures. According to the model temperatures between 15 and 35 °C are conducive to malaria transmission with 25 °C being the optimum for transmission. Calculations were done in Microsoft Excel (Microsoft Inc.).

In the absence of an intervention-free control scenario, R0<0.1 was arbitrarily chosen to delineate areas ‘at risk’ of local malaria transmission.

District-level, monthly malaria case data was obtained from the National Health Information System for the period between 2009 and 2022 (https://www.healthpng.com/).

R_0_ and other spatially explicit results were mapped using Wolfram (Mathematica) 14.2 (Wolfram Research, Inc.). Relationships between R_0_ and other parameters (e.g., altitude, time-of-year)) were plotted in GraphPad Prism 10 (GraphPad Software LLC).

## Results

### Past (1960-1969) to near-present (2010-2019)

Average temperature in PNG, calculated as the average of all grid cells, increased from 23.44°C to 23.75°C between 1960-69 and 2010-19 (ΔT_avg_=0.31 K, 0.1-99.9% centiles: 0.06-0.57 K, Figure 1A and B). The temperature increase was not homogenous across the study area, with southern parts of PNG, in particular Milne Bay Province, being subject to higher ΔT_avg_ (Figure 1A). The temperature increase resulted in subtle changes to the temperature suitability (R_0_) prediction, in particular on the highlands fringes at approximately 850 to 1000 m, where R_0_ increased by an average of 5.6% (range 0.5 to 11.4%, Figure 1 C and D).

**Figure 1:**
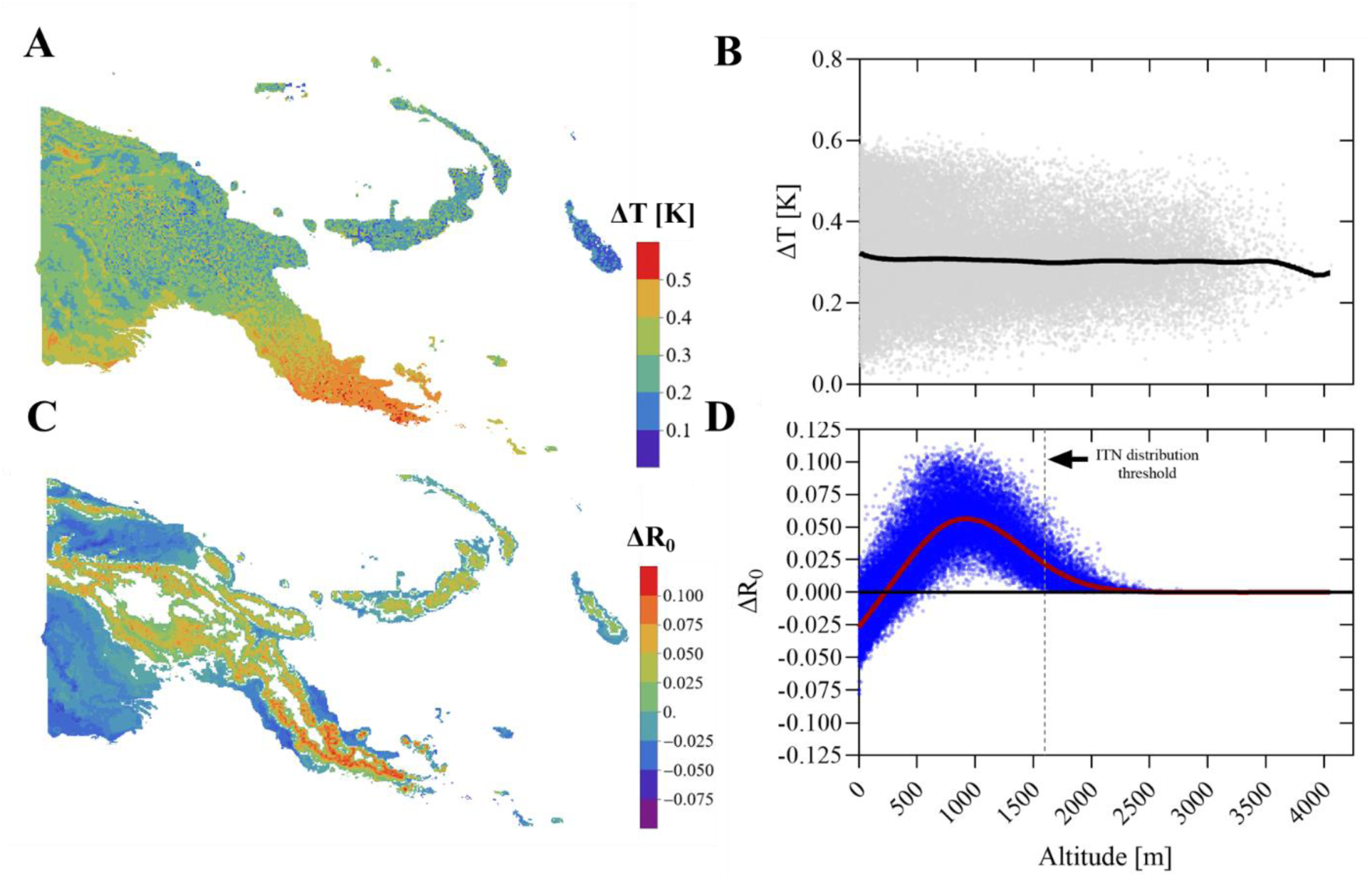
Temperature (T) and temperature suitability (R_0_) change in PNG between 1960 and 2019. Panel A shows the distribution of the average temperature change (ΔT) between the 1960s and 2010s. Panel B: Distribution of ΔT over altitude for each 6.25 square kilometre grid cell between the 1960s and 2010s. Panel C: Change in R_0_ (ΔR_0_) mapped across the landscape. Panel D: ΔR_0_ over altitude including a locally weighted scatterplot smoothing model.

Seasonal temperature fluctuations in PNG are very minimal, and thus had little influence on R_0_ as shown in Figure 2. The ‘seasonal peak’ in ΔR_0_ occurred in October when average ΔR_0_ approached 7.5% at an altitude of about 1000 m. Overall the median altitude limit potentially allowing for malaria transmission shifted by about 46 m altitude.

**Figure 2:**
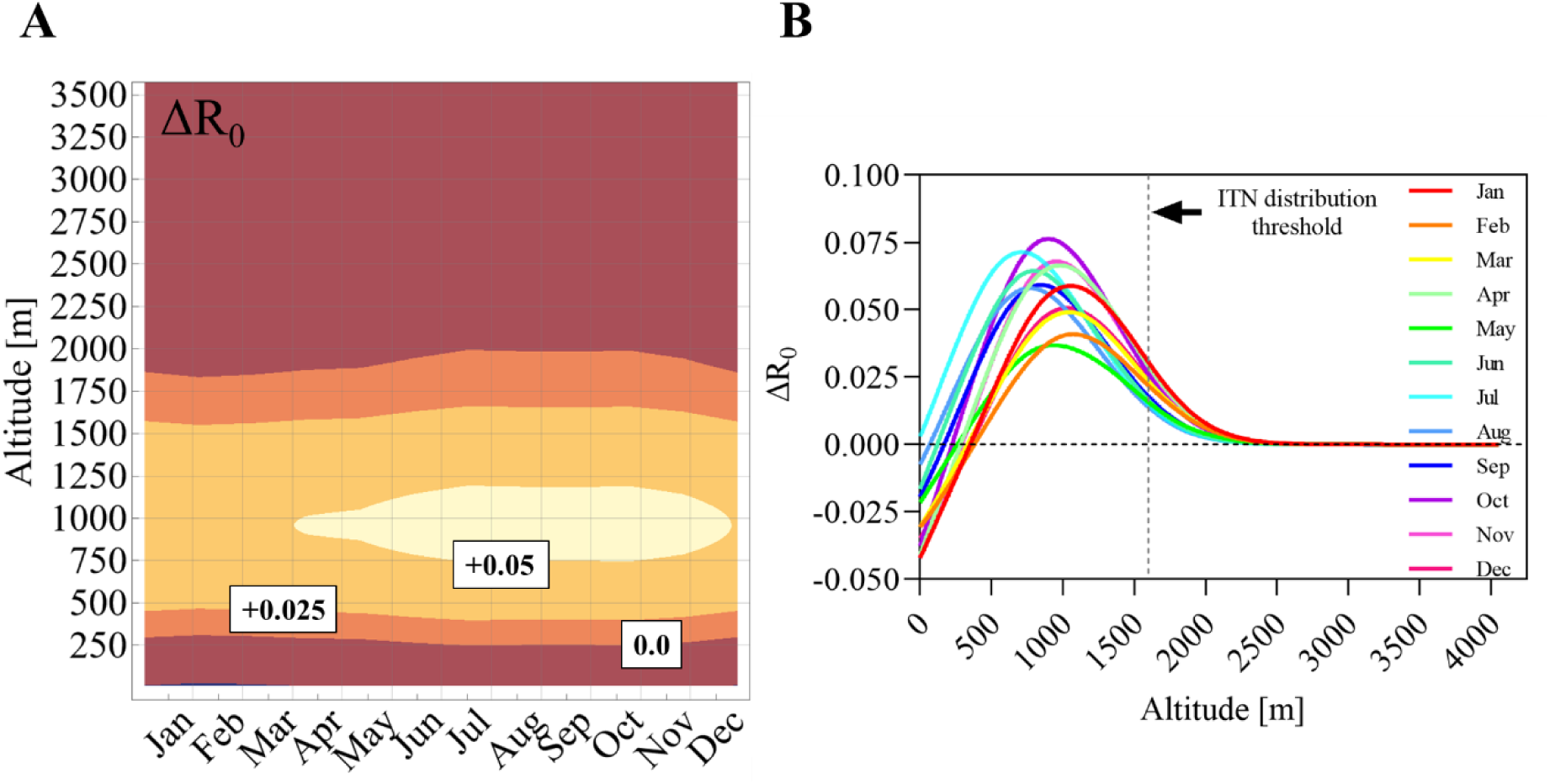
Impact of seasonality and altitude on ΔR_0_ between 1960 and 2019. Panel A: Contour plot showing the monthly average change in R_0_ between 1960-69 and 2010-19 over altitude. The main contours are labelled with the corresponding ΔR_0_. Panel B: ΔR_0_ per month over altitude (one locally weighted scatterplot smoothing model per month). The dashed vertical line is the current altitude threshold for the distribution of insecticide-treated nets (ITNs) in PNG.

### Near-present (2010-2019) to near-future (2021-2040)

Average temperature increased from 23.75°C between 2010-19 to 25.08°C in 2021 to 2040 (ΔT_avg_=1.33 K, 0.1-99.9% centiles: 0.79-1.61 K, Figure 3A and B). The prediction did not follow the same trend as observed with the historical data, which indicated that the southern parts of the country experienced a higher change in temperature. This may have led to a bias in the predicted ΔT distribution between 2010-19 and 2021-40.

**Figure 3:**
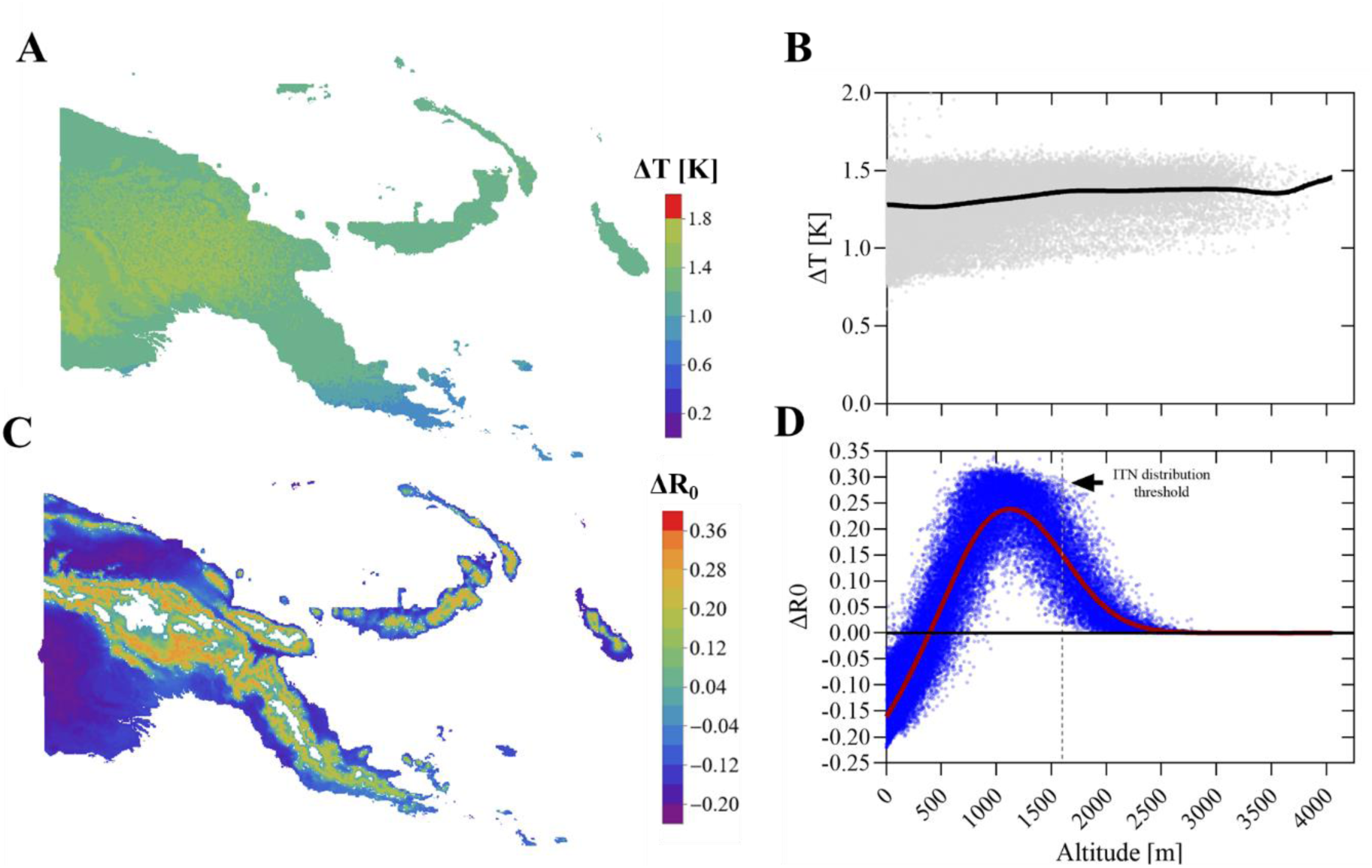
Temperature (T) and temperature suitability (R_0_) change in PNG between 2010-19 and 2021-2040. Panel A shows the distribution of the average temperature change (ΔT) between the 2010s and the 2021-2040 ACCESS CM2 GCM/SSP245 model prediction. Panel B: Distribution of ΔT over altitude for each 6.25 square kilometre grid cell between between the 2010s and the 2021-2040 model prediction. Panel C: Change in R_0_ (ΔR_0_) mapped across the landscape. Panel D: ΔR_0_ over altitude including a locally weighted scatterplot smoothing model.

The projected temperature increase resulted in marked changes to the temperature suitability (R_0_) prediction. ΔR_0_ was highest in the 1000 to 1100 m range, where average R_0_ increased by 23.8% (Figure 3 C and D). Seasonal temperature fluctuations had little impact ΔR_0_ as shown in Figure 4. R_0_ increased by over 20% in some of the most affected altitudes every month of the year. Lower altitude regions below 100 m experienced a drop in R_0_ of 15% or more indicating that conditions may be becoming too hot for optimal malaria transmission. Overall, the median altitude limit potentially allowing for malaria transmission is predicted to shift by 263 m altitude.

**Figure 4:**
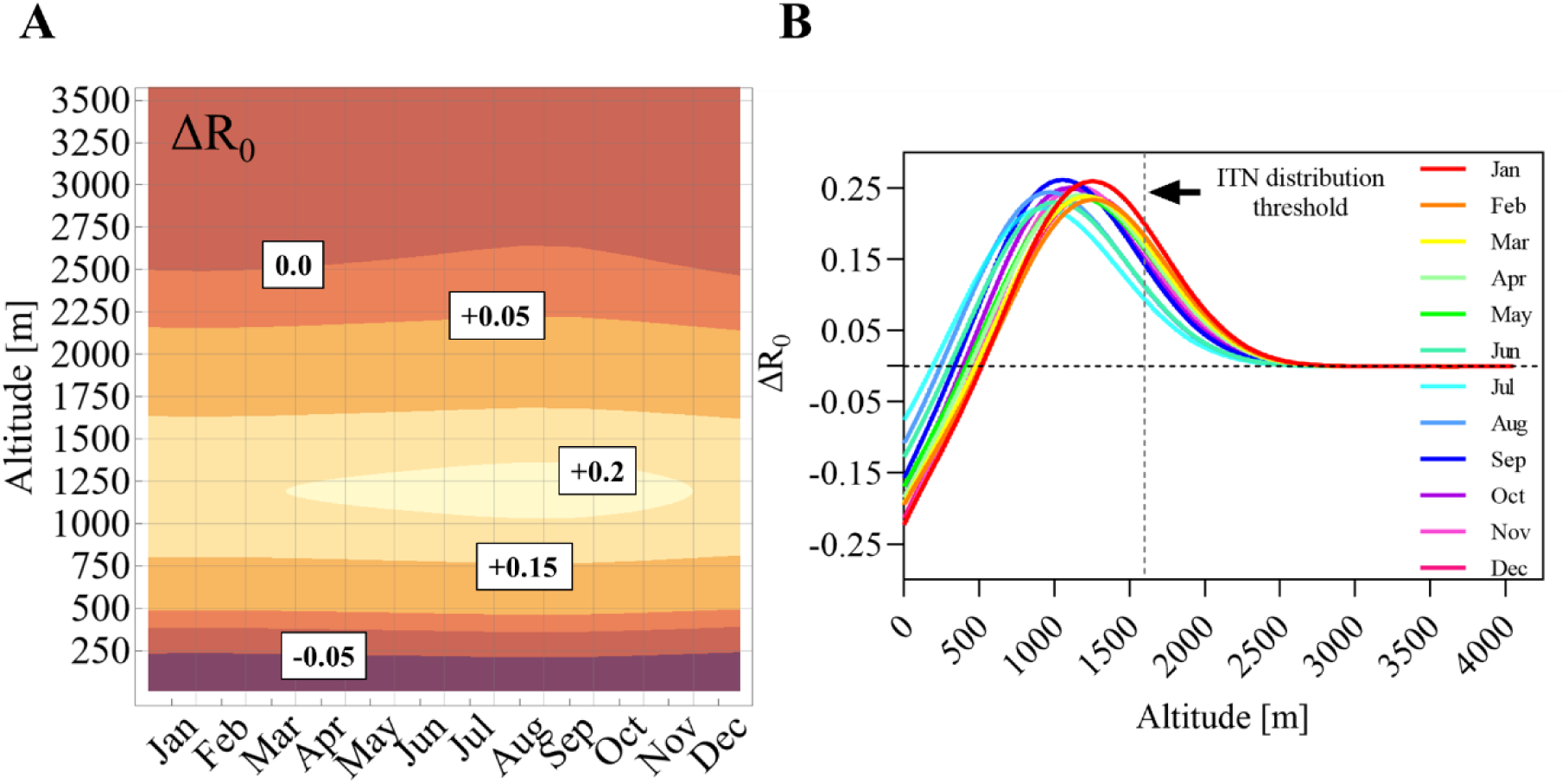
Impact of seasonality on ΔR_0_ between 2021 and 2040. Panel A: Contour plot showing the monthly average ΔR_0_ between 2010-2019 and 2021-40 ACCESS CM2 GCM/SSP245 model prediction over altitude. The main contours are labelled with the corresponding ΔR_0._ Panel B: Average ΔR_0_ per month over altitude (one locally weighted scatterplot smoothing model per month). The dashed vertical line is the current altitude threshold for the distribution of insecticide-treated nets (ITNs).

### Past and future population at risk estimates

PNG population estimates vary considerably according to source. The present study used population estimates from the World Bank (for 1969, https://data.worldbank.org), United Nations Population Fund and the National Statistics Office of PNG for 2019 and 2040 [1, 2]. The population growth rate in PNG was assumed to be 3.1% [2]. The present study used population estimates for the end of each of the study periods, i.e., the years of 1969, 2019 and 2040.

Accordingly, there were an estimated 2,446,927 people in PNG in 1969, 11,244,692 in 2019, and 18,596,012 are projected to live in the country by 2040. Assuming a constant population density distribution across space and time, the cumulative number of people living areas of the country suitable for malaria transmission (as defined by the arbitrary value of an average R_0_>0.1) is plotted over altitude in Figure 6. R0>1 corresponded to median altitude bands of about 1528 m (IQR: 1404-1632m) in 1960-69, 1574 m (IQR: 1458 1686m) in 2010-19 and 1837m (IQR: 1729 to 1944m) in 2021-2040.

**Figure 5:**
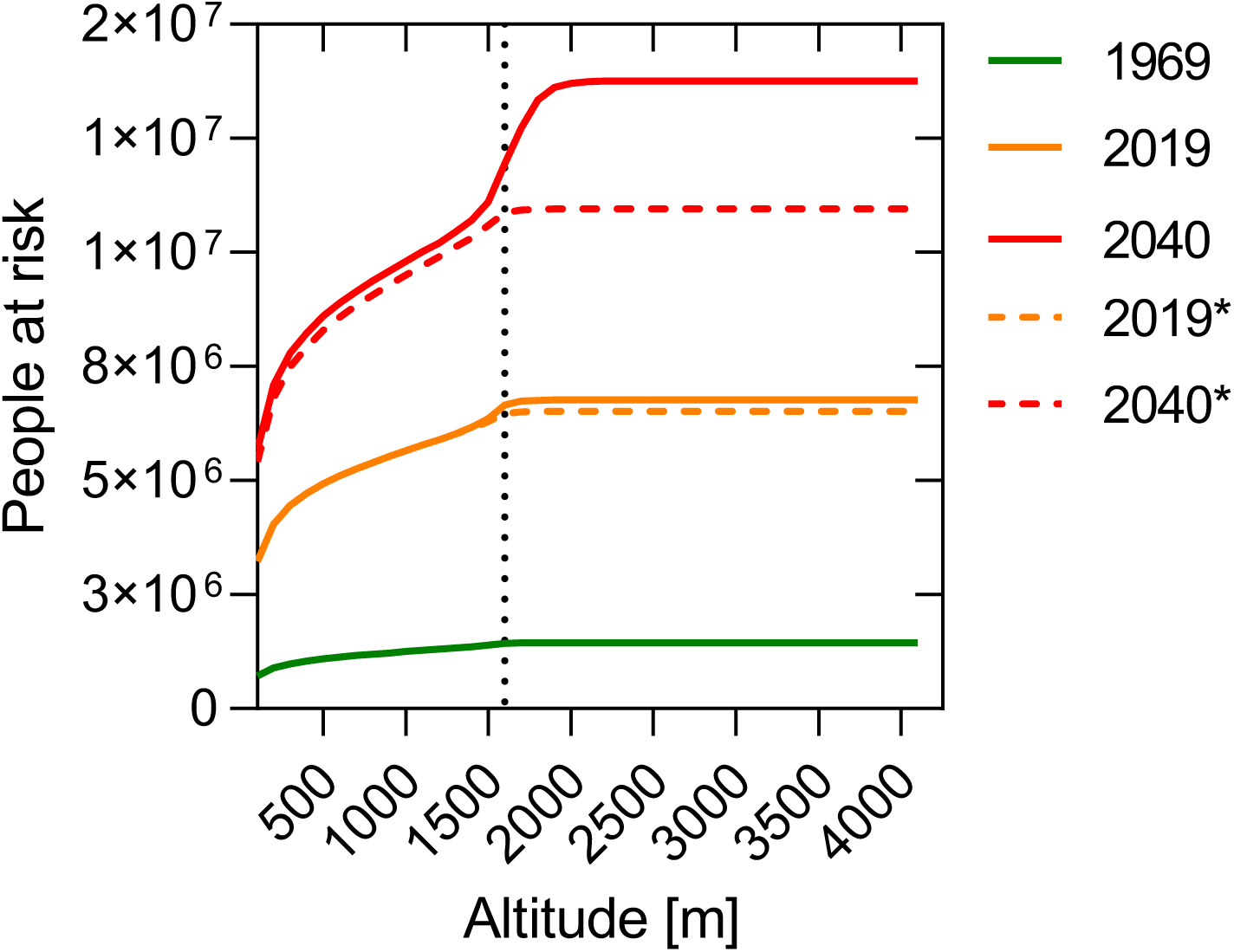
Cumulative population at risk estimates over altitude in 1969, 2019 and 2040. Solid lines are estimates accounting for population growth and climate change. Dashed lines (labelled *) are estimates accounting only for population growth (i.e., using the R_0_ estimates of the 1960-69 period). The vertical dotted line is the current ITN distribution altitude threshold.

**Figure 6:**
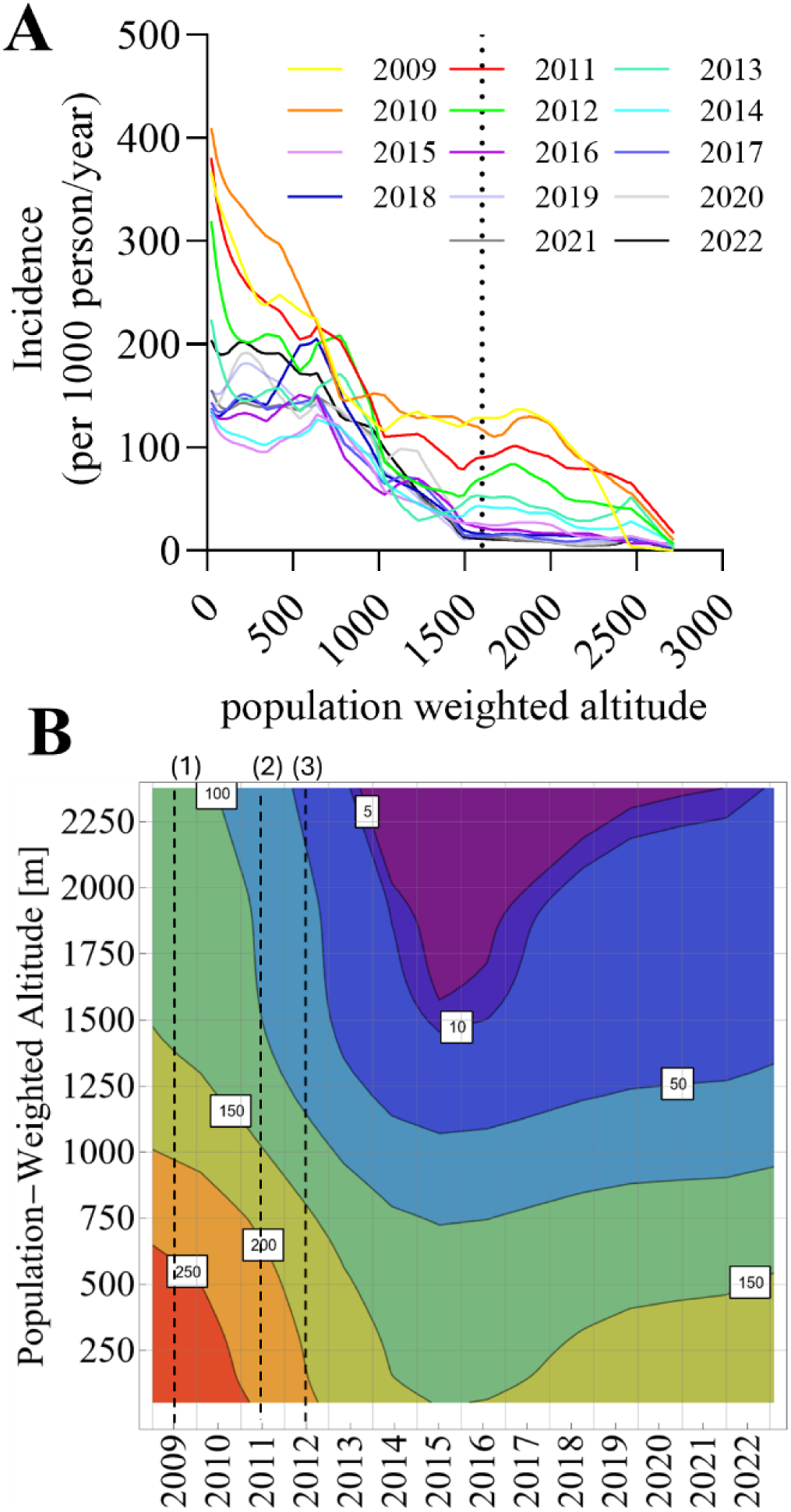
Relationship of malaria incidence, time and altitude in PNG. Panel A) Average malaria incidence over population-weighted altitude for years 2009 to 2022. Averages were calculated using monthly incidence data for each PNG district and applying a LOWESS model (raw data not shown). Panel B) Contour plot visualising the relationship between year, altitude and malaria incidence (contours). Data was smoothened using a generalised additive model with 5^th^ order polynomial splines. The dashed lines denote the introduction of ITNs (1), ACTs (2) and RDTs (3).

Figure 6 also shows scenarios where population growth, but not climate change, was taken into account. While in 1960, about 59% of the total population lived in areas of PNG with temperature suitability >0.1, the proportion increased to 61% in 2019, and is expected to increase to 74% in 2040.

The number of additional people living in areas suitable for malaria transmission due its range expansion was 249,125 in 2019 and is expected to increase to 2,802,709 by 2040.

Overall, the number of people living in areas in areas suitable for malaria transmission in PNG was estimated to 1.44m in 1969, 6.77m in 2019 and 13.76 in 2040. Thus, the resources needed to maintain the current control effort can be expected to more than double by 2040.

### Limits of Current Intervention Effectiveness and Populations at Risk of Ineffective Malaria Control

Interventions impact malaria transmission on much shorter timescales than climate change. However, the degree by which interventions reduce malaria incidence may also depend on how stable the transmission environment is [21].

The 2010 to 2019 decade saw a significant upscaling in malaria control interventions in PNG [22]. Specifically, ITNs are being mass-distributed since 2009, rapid diagnostic tests are being scaled up since 2012, and Artemisinin Combination Therapy was recommended in 2011, i.e., three important interventions were introduced nearly at the same time [23-25]. Programmatic malaria surveillance data, summarised in Figure 7, indicate that these interventions coincided with a disproportionally greater reduction of malaria incidence in highlands areas, where transmission is less stable. More specifically, malaria incidence in most areas over 1500 m reduced by over 10-fold between 2009 and 2022, whereas malaria incidence in areas below 500 m was similar in 2022 as it was in 2009. And while malaria incidence in coastal areas is resurging since 2015, this is not indicated for the highlands. Thus, while higher altitude regions may become more suitable to sustain transmission due to climate change, current interventions may compensate for some of the impact.

**Figure 7:**
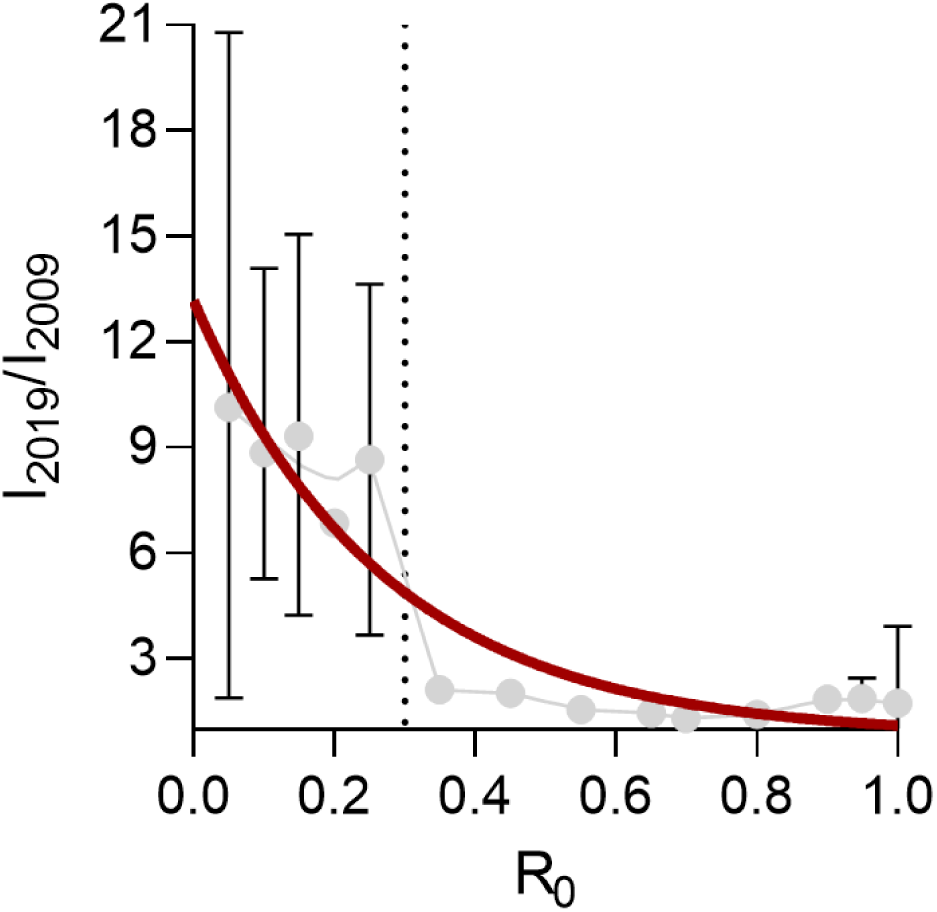
Fold-decrease of malaria incidence between 2009 and 2019 versus R0. I_2009_ and I_2019_ denote the incidence in 2009 and 2019, respectively. The error bars are the ranges of incidence observations within R_0_ +/- 0.025 increments. The red curve is a guide to the eye (simple exponential decay model). The dotted vertical line is R_0_=0.3, which appears to be an adequate threshold to delineate current intervention effectiveness.

To estimate this compensatory effect, the fold-decrease in malaria incidence in each district (n=82) within the 2009-2019 decade is plotted over the corresponding R_0_ for the same decade in Figure 8.

**Figure 8:**
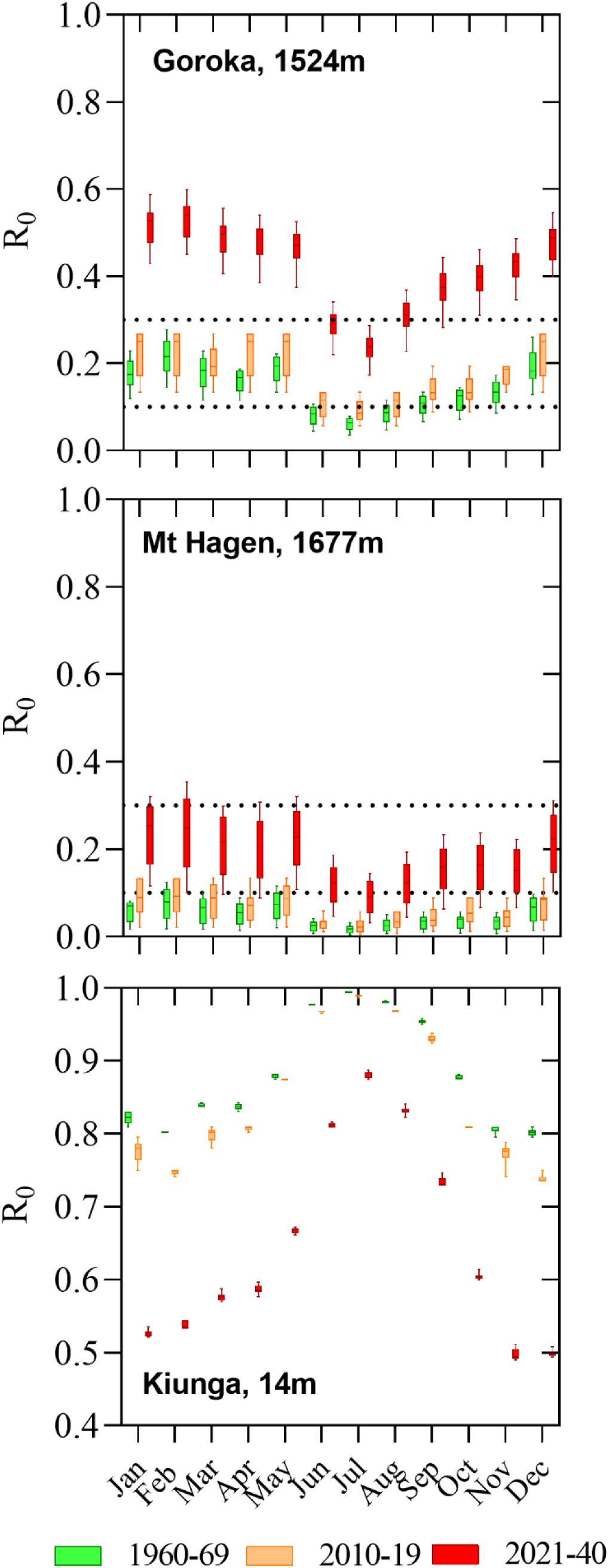
Examples for temperature suitability changes in PNG population centres located at different altitudes. Panel A: Goroka (Eastern Highlands Province, 1524m); Panel B: Mount Hagen (Western Highlands Province, 1677m); Panel C: Kiunga (Western Province, 14m). The box and whisker charts summarise the data for all grid squares belonging to the specific local-level government (LLG) area. Data for the highlands LLGs appears more spread due to the larger differences in altitude within these LLGs, as compared to Kiunga. The dotted lines denote R_0_=0.1 (arbitrary cut of denoting malaria risk areas) and R_0_=0.3 (malaria stable accounting for current interventions).

The analysis indicated that the decrease in reported malaria incidence was much more pronounced in areas of R_0_<0.3. Based on this, a R_0_ threshold was set, R_0_=0.3, denoting the limit of effective malaria control being achieved with the current set of interventions. This corresponds to a median altitude of 1230 m in 2019 and 1519 m in 2040. When this threshold was applied to calculate the populations living in areas where the current set of interventions may not result in a further reduction of incidence, the resulting numbers were 5.92 million in 2019 and 11.40 million in 2040. Climate change accounted for 1.43 million (26%) of the estimated increase between 2019 and 2040, while population growth accounted for the remainder (74%).

### Specific Example Population Centres

To illustrate the changing temperature suitability for malaria transmission, three specific example local level government areas (LLGs) were selected, a) the two highlands population centres of Goroka (Eastern Highlands, 1524 m) and Mount Hagen (Western Highlands, 1677 m), and b) the lowland population centre of Kiunga (Western Province, 14 m). Figure 5 shows monthly ranges R_0_ for these LLGs for the three time periods 1960-69, 2010-19 and 2021-40. In the highlands population centres, R_0_ did not change much between 1960-69 and 2010-19, but temperature suitability is expected permit year-round malaria transmission in 2040 and, in case of Goroka, is predicted to be in the range of R_0_>0.3 where current interventions are not able to reduce malaria incidence further. In Kiunga, the observed trend was inverse, and temperatures are predicted to become too hot to sustain optimal malaria transmission by 2040. R_0_ in the Kiunga scenario decreases in particular in the southern hemisphere summer months.

## Discussion

The present study aimed to estimate the impact of climate change on malaria transmission temperature suitability in PNG, which currently reports the highest malaria incidence outside of Africa [4]. Quantifying the effect of climate change to the region’s malaria transmission is cruicial in the context of malaria elimination, by which nations including PNG have committed to eliminate malaria in the near future [14]. Projections such as those from the present study can help to more accurately estimate future resource needs for malaria control and prevent under-resourcing.

Our findings estimate that by 2040, 7 million additional people in PNG will live in areas at risk of malaria transmission, with 2.8 million of these cases directly attributable to climate change and the remainder due to population growth. Consequently, the PNG malaria control program may need to distribute an additional 1.2 million insecticide-treated nets per year — double the current number — just to maintain existing control efforts.

Temperature plays a crucial role in determining malaria transmission suitability, serving as a fundamental requirement for an area to sustain malaria transmission [26, 27]. While other factors, such as the availability of larval habitats for competent vectors, also shape real-world transmission intensity [28], temperature sets the baseline conditions necessary for vector and parasite development. Many of these factors, including precipitation and land-use, are also impacted by climate change and anthropogenic influence [29].

The vectors involved in malaria transmission in the PNG Highlands and their habitats are still very poorly described. Several studies have reported *Anopheles farauti* 6 and *Anopheles bancroftii,* and *Anopheles punctulatus s.s*., *Anopheles kawari* and *Anophlees sticmaticus* have also been found [30-32]. There is an urgent need for systematic vector surveillance data in these changing ecologies to inform interventions and models.

Malaria epidemics can occur in the PNG Highlands, and have been observed in most highland provinces, in villages usually at altitudes of 1400 to 1700 metres, in particular prior to the implementation of ITNs, RDTs and ACTs [33-37]. The malaria prevalence during these epidemics can exceed 30% of the population, with significant clinical disease [33]. The most recent epidemic was reported in (2022) with >250 cases reported and many more likely unreported [38]. While there is no analysis of the frequency of epidemics over recent decades, they appear to have become less frequent, likely as a result of the disproportionate impact of interventions in the highlands areas also described in this study.

Despite the growing concern over climate change, research on its impact on malaria transmission in PNG remains limited. One study linked increased malaria incidence in the Eastern Highlands Province to climate change between 1996 and 2008, prior to the widespread implementation of several new interventions [39]. A school survey from 2019 reported low malaria prevalence among PNG Highland school children, with infections predominantly caused by *P. falciparum* [40]. However, the 2022/2023 malaria indicator survey found surprisingly high malaria prevalence in several high-altitude villages, with 30% RDT positively reported in two villages above 2000m in Enga Province and an 8.6% average *P. falciparum* prevalence across three villages betweem 1500m and 2000m in Chimbu Province including children under 5 [41]. These findings suggest local transmission in areas previously considered unsuitable for malaria, potentially signaling an ongoing epidemic. Further research and surveillance capacity strengthening is needed to monitor malaria incidence in PNG more accurately in particular with ongoing interventions. However, resource limitations and difficult access represent immense challenges.

The analyses presented here assumed that population density distribution in PNG remained constant over the study period. However, this assumption is unlikely to reflect reality as urbanisation and internal migration, - which are also impacted by climate change – continue to reshape population patterns [42]. Understanding how these demographic shifts intersect with changing malaria transmission zones is essential. There is a need for more research quantifying internal human migration in PNG and to assess how population movement may influence malaria risk and control efforts.

The analysis relied on previously established parameters, originally developed for African malaria vectors (in colony) and *Plasmodium falciparum* [15]. Some details regarding the generalizability of these estimates have been challenged by other malaria modelling groups [28]. In general, it should be acknowledged, that vector and parasite species and strains vary in terms of these parameters, and that PNG malaria vectors and parasites, in particular *Plasmodium vivax,* exhibit locally specific characteristics that are likely to modulate the R_0_(T) curve. For example, evidence suggests that *Plasmodium vivax* undergoes extrinsic development in the mosquito more quickly as compared to *P. falciparum* at the same temperature [43]. Yet, while *P. falciparum* accounts for at least 75% of reported cases in PNG, there is evidence that *P. vivax* may be disproportionately prevalent in the higher altitude regions due to its resilience against interventions and its propensity to survive in colder climates [7]. Most previous surveys have relied on light microscopy and RDTs, which may fail to detect the majority of *P. vivax* infections [40, 44]. To refine current models and better capture malaria dynamics, experimental research is needed to establish temperature-dependent development profiles for with PNG’s vectors and parasites. Expanding the present analysis to include *P. vivax* temperature suitability ranges will be crucial for developing more accurate projections of malaria transmission risk under future climate scenarios.

Climate projections are based on complex General Circulation Models (GCM) and Shared Socioeconomic Pathway Models (SSP), each making distinct assumptions about future climate conditions [45]. The present study used athe ACCESS CM2 GCM, developed in Australia (https://research.csiro.au/access/about/cm2/) and one SSP (SSP245) model combination. This can be considered a conservative scenario where climate change is mitigated well, CO_2_ emissions peak in the 2040-49 decade, and overall temperature increase is limited to the 1.5–4.5 °C range by 2100 [46]. However, the likelihood of achieving this scenario is diminishing as global emission trends suggest more severe outcomes may be likely., To strengthen future projections, it may be imporant to explore different model combinations, and more pessimistic scenarios predict.

The present study made simple assumptions about populations at risk and resource needs. The cut-off of R_0_ (T_avg_) > 0.1 is only a crude estimate to delineate at-risk areas, which coincides with the current ITN distribution threshold. Seasonality and other factors are likely to further shape the true relationship of temperature and malaria transmission. Furthermore, it may be that malaria naïve highlands populations are much more susceptible to clinical disease and severe *falciparum* malaria than their coastal counterparts, due to less immunity [7]. This could potentially increase resource needs significantly and non-linearly, especially those for diagnostic tests and treatments, which is not considered in our model estimates. Conversely, the lack of immunity may increase the frequency of treatment seeking and thus reduce transmission by asymptomatic carriers [47]. It is therefore important to develop more detailed models to more accurately predict these resource needs.

Interventions had a significantly different impact on malaria incidence across R_0_ strata in the 2010 to 2019 decade and generally appear to have been much more effective in the highlands. This is expected, as transmission chains are more easily interrupted where the transmission environment is unstable (i.e., R_0_ is low) [48]. The present analysis indicated that the cut off between effectiveness and non-effectiveness to sustainably reduce malaria incidence of the current malaria control effort in PNG is around R_0_=0.3. This corresponds a median altitude range of 1230 m (IQR: 1111-1349m) in 2019 and 1519 m (IQR 1415-1614 m) in 2040. Thus, in current and future areas with R_0_>0.3, the presently applied intervention mix is unlikely to result in sustained reduction of malaria incidence. This results in multiple challenges for the PNG malaria control program i) to expand the current intervention mix into areas that are becoming suitable to malaria transmission due to climate change, to maintain current gains, and prevent resurgence and ii) to implement new interventions that address current protection gaps [49] and bring down malaria rates further.

A resurgence of malaria in the PNG Highlands would not be without precedence, as this occurred after the collapse of the control effort in the early1980s [50]. In line with several previous publications, this work provides further evidence that without significant additional resources, malaria elimination in PNG will become more elusive instead of more tangible in the near future, in contrast to current regional elimination goals.

## Conclusion

Climate change is expected to shift the temperature suitability range in PNG towards higher altitudes, expanding the population at risk and complicating malaria control efforts. This study highlights the critical need to account for climate-driven shifts when planning for future malaria elimination strategies. We estimate that by 2040 an additional 7 million people may live in areas suitable for malaria transmission, with 2.8 million directly attributable to climate change. To prevent a resurgence of malaria, as seen in the PNG Highlands after control efforts collapsed in the 1980s, multiple challenges must be addressed; expanding current interventions into newly at-risk areas, maintaining gains in regions with unstable transmission and introducing new strategies to close protection gaps. More research is needed to accurately quantify additional resource needs to achieve effective malaria control and elimination and to better understand the biological interactions between temperature and PNG malaria vectors and parasites.

## Data Availability

All data produced in the present study are available upon reasonable request to the authors

## Acknowledgments

We thank the PNG National Malaria Control Program for support.

